# Incidence and clinical predictors of Continuous Positive Airway Pressure (CPAP) failure among preterm neonates: a prospective clinical research study protocol

**DOI:** 10.64898/2026.05.20.26353688

**Authors:** Rhobi Gabriel Sisa, Salvatore Florence Kalabamu, Maulidi Rashidi Fataki, Nasreen Ayoub Daud, Ansaar Ismail Sangey, Kelvin Melkizedeck Leshabari

## Abstract

**Introduction:** Newborn babies frequently encounter acute respiratory failure requiring assisted ventilation. Acute respiratory failure in infants commonly present in a form of respiratory distress syndrome. There are several studies that documented factors associated with CPAP failure rates among preterm newborns worldwide. However, they were either statistically underpowered or defined by overt design errors. The proposed study will estimate incidence rate and predictors of Continuous Positive Airway Pressure (CPAP) failure among preterm newborns delivered at representative hospitals in a typical urban area of Africa.

**Methods and analysis:** a prospective longitudinal cohort observational, analytical study will be conducted at neonatal and emergency units of all Dar es Salaam public regional referral hospitals from March to (and including) August 2026. Newborns with CPAP failure will be the target population. Newborns without CPAP failure will be the control group. Follow-up for each child will commence from the moment the child is subjected to CPAP until CPAP failure is clinically evident or day seven of life, whichever comes first. Interval assessment of the SAS scores (for CPAP potency) will be done using Silverman-Anderson score sheet in 4-6 hours intervals (unless otherwise dictated by the child’s clinical situation). The main outcome/dependent variable will be proportion of new CPAP failure per newborn-time of follow-up. A multivariable linear model will be used to account for independent predictors of CPAP failure. Unless otherwise stated, an α-level of 5% will be used as a limit of type 1 error in findings.

**Ethics and Dissemination process:** The study has received an IRB certificate (IRB reference: KU/IREC/27.10/639) from the Institutional Research Ethics Committee of KU. Permission to recruit the affected children has been sought from Municipal’s based hospitals’ directors of Amana, Mwananyamala and Temeke regional referral hospitals respectively. Written informed consent will be sought from mothers of all recruited newborn babies.

**Strengths and limitations of this study:**

+ The findings of this study will be probably the first original findings to characterize incidence rate and clinical predictors of CPAP failure among newborns according to their birth stata (extreme preterms vs preterms vs terms vs. >week 42 newborns)
+ The study findings will bring about the incidence rate measure, unlike others in the past that mainly reported their burden using prevalence estimates.
  - Referral biases may not be adequately controlled by this follow-up study since all our recruits will be from referral hospitals with both NICU and emergency facilities for acute respiratory failure in newborns.
  - Potentially high attrition (due to say early deaths?) rates among followed-up newborns.

## Background

### Context

According to UN estimates, about 2.3 million newborn children died globally in 2024 alone – equivalent to about 6,200 newborn deaths per day [1, 2]. The statistics reflect age-standardized death rate of around 17.2 per 1000 live birth worldwide [1]. Africa is the most affected region of all at global scale [1-3]. By far, infectious diseases like pneumonia, diarrhoeal diseases, malaria as well as preterm birth complications; birth asphyxia, trauma and congenital anomalies remain the leading causes of deaths among newborn babies worldwide [3]. Even in Africa, respiratory failure and its associated conditions (birth asphyxia, respiratory distress syndrome, meconium aspiration pneumonia – for newborns; pneumonia and febrile illnesses– for under-five years) have been documented to be the chief culprit for under-fives deaths [4-7]. Amidst all those frustrating statistics, there is still paucity of data about the exact contribution of its individual precursors, especially prematurity and respiratory distress syndrome. Recent analysis reported from Dar es Salaam in Tanzania revealed of the 128 newborn babies born with prematurity, 35.9% (n= 46) died [5]. Similarly, in the same study - of the 43 newborn babies with established respiratory distress syndrome, about two-thirds died within the first seven days of life [5]. At present, there are compelling indications that suggest interventions tailored to newborn with prematurity and respiratory distress syndrome increase survival chances [8-11]. However, little has been established about nature of the standard interventions strategies for newborns with respiratory distress in developing nations – Africa inclusive. Besides, there are also a body of literature that narrates about failure rates for the same survival-based intervention strategies [12, 13]. The proposed study will estimate incidence rate and predictors of Continuous Positive Airway Pressure (CPAP) failure among preterm newborns delivered at representative Dar es Salaam hospitals.

Premature newborns frequently encounter acute respiratory failure requiring assisted ventilation [14, 15]. By far, acute respiratory failure (in form of Respiratory Distress Syndrome – RDS) forms one of the commonest indications for neonatal intensive care unit admission worldwide [14, 16-18]. Respiratory failure refers to inability of body maintenance either towards normal delivery of oxygen to tissues or optimal expulsion of carbon dioxide from tissues [19]. Technically, the imbalance between respiratory workload and ventilatory strength and endurance precisely defines respiratory failure, be it among newborns or adults. Even though the definition of respiratory failure is somewhat arbitrary, it is objectively defined using laboratory and clinical criteria that includes oxygen saturation < 80% or PaO_2_ < 50mmHg, PaCO_2_>60mmHg with an FiO_2_ of 1.0 and pH<7.25 [20]. In infants and younger humans, acute respiratory failure commonly present in a form of respiratory distress syndrome. Respiratory distress syndrome in newborns develops due to immaturity of surfactant synthesis system, insufficiency of surfactant production and/or structural immaturity of the lungs [20].

Previously, standard treatment for premature infants’ respiratory support included assisted ventilation and surfactant [21, 22]. However, evidence accumulated that assisted ventilation in extremely younger humans damages the lungs [23-25]. Specifically, bronchopulmonary dysplasia ranks among the most acknowledged complications of assisted ventilation among premature infants worldwide [26-28]. Recently, RCT findings from different countries and continents all together – both individually [15, 16, 18] and collectively [18, 27], suggest that initiation of nasal continuous positive airway pressure to be non-inferior (if not superior) to intubation and/or surfactant administration among premature newborns [15, 16, 18, 27]. Conversely, usage of mechanical intubation is at present reserved to those newborns with acute respiratory failure, whose CPAP measures have failed. There are several studies that documented factors associated with CPAP failure rates among preterm newborns worldwide [15, 29-32]. However, they were either statistically underpowered [15] or defined with overt design errors [29-32]. For instance, large sample analysis (N= 19103 newborns; CPAP-administered = 11684) findings from Australia and New Zealand NICUs dataset revealed CPAP failure rates of 43% (n= 863) and 21% (n= 2061) for infants commencing on CPAP at 25-28 and 29-32 weeks’ gestation cohorts respectively [31]. CPAP failure was significantly associated with bronchopulmonary dysplasia (A.O.R.: 1.94, 95% C.I.: 1.44 – 2.61 [25-28 weeks old newborns] and 3.32, 95% C.I.: 2.51-4.39 [29-32 weeks old newborns]) and deaths (2.82, 95% C.I.: 1.4 – 5.65 [25-28 weeks old newborns] and 6.04, 95% C.I.: 2.47 – 14.8 [29-32 weeks old newborns]) [31]. However, the entire analysis relied on reported data from network registry [31]. The registry had restriction to some of the detailed clinical, associated-risks as well as imprecision associated with adjusting for risk profile within the study groups that were also acknowledged by investigators in their published report [31]. Besides, it is unclear whether those findings could be representative worldwide. Thus, the need for fresh data is justified.

### Current knowledge

Continuous Positive Airway Pressure (CPAP) failure in neonates refers to the inability to maintain adequate respiratory function with CPAP support, necessitating escalation to invasive mechanical ventilation or other advanced respiratory interventions. Current evidence indicates that failure may occur in 20-40% of neonates started on CPAP, which necessitates intubation and mechanical ventilation [30]. This raises the mortality risks, intraventricular haemorrhage and bronchopulmonary dysplasia BPD [33, 34]. The criteria for CPAP failure include; persistent or worsening respiratory distress (Increased respiratory rate (>70 breaths per minute), severe retractions, grunting, nasal flaring, or apnoea; Downes score >6 or Silverman-Anderson score >6 despite CPAP at 6 cm H_2_O) [35]. FiO_2_ requirement > 0.38 to 0.6 for > 30–60 minutes to maintain SpO_2_ 90–95%; ≥3 apnoea episodes requiring stimulation in an hour, any episode requiring bag-mask ventilation despite CPAP support, poor perfusion, prolonged capillary refill, or bradycardia not related to apnoea [35]. A multi-centre prospective observational study conducted in 46 neonatal units in France, showed a failure rate of 12% which was defined as the need for tracheal intubation 72 hours after CPAP initiation [34]. In their findings the key predictors were singleton pregnancies, lower Apgar score at 10 minutes, lower gestational age, higher FiO2, surfactant use within 24 hours of life[34]. However, the study findings were derived exclusively from a post-hoc analysis, and exclusively at a referral centre in a developed nation, with reported limited data of late preterm infants; and hence its findings are unlikely to be generalizable not only in France but also in developing nations, where CPAP failure is even more pronounced.

In Africa, there are several reported research reports on the concept of CPAP failure and its associated factors (12, 36-38). However, they are all characterized by design, analysis or reporting defects (12, 36-38). For instance, Abdallah and his colleagues reported their prospective observational findings from a national hospital in Dar es Salaam, Tanzania that revealed overall CPAP failure rate of 37.4%, with most CPAP failure occurring within the first 24 hours (12). However, the study was significantly underpowered to detect CPAP failure (12). It was not surprising, therefore – they could not find any independently factor associated with CPAP failure int heir report (12). For the rest of reported studies in Africa, they had overt errors in analysis and even reporting of their findings to warrant significant attention in this agenda (36-38).

## Evidence and population gap

A noticeable proportion of neonates still experience CPAP failure characterized by persistent or worsening of respiratory distress (grunting, lower chest-wall indrawing, nasal flaring) despite Continuous Positive Airway Pressure (CPAP) proven efficient at raising oxygen levels and lowering demands for invasive mechanical ventilation among newborns (15, 29-32). In particular, there appears to be evidence (12, 34, 35) and knowledge gaps (34) in the reliable estimates of CPAP failure in newborns with respiratory distress syndromes. In particular, of special interest to clinician-researchers as well as neonatologists the world over, are the clinical predictors of CPAP failure among newborns, especially those born preterm. Moreover, of even special interest is to analyse if there are any significant differential incidence and predictors among those born preterm from those born at term to those who are born after 42 weeks of gestation process.

## Objectives

### Broad objective

To determine the magnitude and clinical predictors of continuous positive airway pressure failure among preterm neonates diagnosed with respiratory distress syndrome.

### Specific objectives

1. To determine the incidence rate of CPAP failure among neonates diagnosed with respiratory distress syndrome.
2. To determine clinical predictors of CPAP failure among preterm neonates with respiratory distress syndrome.

## Rationale

Understanding the magnitude and clinical predictors of CPAP failure among newborns have several benefits, especially in lieu of the fact that newborns respiratory distress syndrome is among key causes of newborn deaths worldwide (1, 4-7). Improvement in global under-five deaths statistics in recent years has been slowed by disproportionate higher levels of newborn deaths, especially in Tanzania (39).

Moreover, there is also palpable evidence that suggest early life respiratory illnesses to be precursors for morbid and increased mortality risks later in life worldwide (41-44). This becomes handy the world over now, since demographic transition process has been widespread, even in Africa (45). Tanzania in particular, is an East African nation with the fastest reported demographic transition process (45). Besides, there are incoming evidence that suggest increased healthcare spendings and outcomes associated with chronic diseases among senior citizens, almost always with precursors early on in life (46-50). At local context, there is a new wave of scientific inquiry that endeavours to build evidence in terms of measurable outcomes across the entire human lifespan (51-54). Thus, the rationale for the current epidemiological analysis.

## Methods

### Study design

The proposed study protocol will be a prospective longitudinal cohort observational, analytical study. The decision to such design reflected the dire need for observational follow-ups of each studied preterm with respiratory distress syndrome for a maximum duration of seven days post-delivery. Those who will ultimately fail on CPAP will be the exposed group. Those who will survive on CPAP will be the non-exposed group. Follow-up for each child will commence from the moment the child is subjected to CPAP until CPAP failure is clinically evident or seven days of life whichever comes first. Interval assessment of the SAS scores (for CPAP potency) will be done using Silverman-Anderson score sheet in 4-6 hours intervals (unless otherwise dictated by the child’s clinical situation) per protocol.

### Study settings

Amana, Mwananyamala and Temeke regional referral hospitals in Dar es Salaam. Those three hospitals are the only public regional referral hospitals in Dar es Salaam metropolitan city. Whereas Amana hospitals has 15 neonatal beds with five neonatal CPAP machines receiving delivered and referred neonates primarily from Ilala municipality, Mwananyamala has 25 neonatal beds with seven neonatal CPAP machines receiving delivered and referred newborns from Kinondoni and Ubungo municipalities. Temeke regional referral hospital has 20 newborn beds with five neonatal CPAP machines receiving delivered and referred newborns from within Temeke and Kigamboni municipalities.

### Follow-up period

Six months’ span from March to August 2026.

### Sampling method

All preterm neonates delivered to the three (Amana, Mwananyamala and Temeke) public regional referral hospitals in Dar es Salaam city admitted with respiratory distress syndrome and initiated on CPAP in the hospitals’ neonatal units will be enrolled sequentially as they present until the required sample size of is attained meeting the inclusion criteria provided that they meet the eligibility criteria. This method was chosen because of the maximization of study participants with limited number of neonatal beds and neonatal CPAP facilities. Otherwise, the benefit also includes the fact that no sampling biases/errors prevalent in other quantitative sampling methods. Eligibility criteria will include preterm (age≤37 weeks) neonates delivered within the past six hours, symptoms/signs of respiratory distress syndrome and hospitals’ protocol need for CPAP initiation. All newborns with congenital anomalies and those who will be referred to higher referral facilities (i.e. Muhimbili National Hospital) as well as those with respiratory distress syndrome not requiring CPAP (e.g. those with low or high flow nasal cannula oxygen therapy) will be excluded from this follow-up study.

A minimum sample size of 196 newborn babies with respiratory distress syndrome in need of CPAP therapy will be needed to answer the primary research question (i.e. *what is the incidence of CPAP failure among newborns-time of follow-up at Dar es Salaam hospitals*?) given apriori 5% level of significance for detection of type 1 error and considering study power of at least 80%.

### Study variables

The main outcome/dependent variable will be proportion of new CPAP failure per newborn-time of follow-up which is defined by the occurrence of clinical deterioration requiring escalation of respiratory support based on criteria such as: FiO2>60% for more than one hour, three or more episodes of apnoea requiring stimulation within an hour, bradycardia (heart rate < 100 beats per minute measured by pulse oximeters), poor perfusion (capillary refill >3 seconds) or the need for bag-mask ventilation.

Besides, a number of independent/predictor variables will include a range of neonatal and maternal factors such as: gestational age in weeks (to be treated as *a continuous-interval variable*), birth weight in kilogrammes (*a continuous-interval variable*), gender (binary variable of 1-female vs 2-male), Apgar scores at 1 and 5 minutes (*continuous-interval variables*), mode of delivery (categorical variable). Clinical indicators such as Silverman-Anderson scores (a continuous-interval variable) and initial SPO2 (*a continuous-interval variable*), initial FiO2 (a continuous-interval variable), CPAP pressure settings (*a continuous-interval variable*), the time of CPAP initiation (a continuous-interval variable), number of apnoea episodes (*discrete variable*), presence of bradycardia (binary variable of 1 – present vs. 2 - absent) and requirement for bag-mask ventilation (binary variable of 1 – present vs. 2-absent). Maternal and perinatal variables such as antenatal corticosteroids use (binary variable), place of delivery (categorical variable) and multiple gestations (discrete variable) will also be included.

### Data collection procedure

Tools used to collect data will be Case Report Forms (CRFs) specifically designed for this study to capture comprehensive neonatal, maternal, and clinical data. These forms will include information on gestational age, birth weight, gender, mode of delivery, APGAR scores, antenatal corticosteroids use, maternal infections and place of delivery. Clinical indicators such as Silverman-Anderson Score, FiO2 requirements and signs of respiratory distress will be recorded at baseline and every 4-6 hours after CPAP initiation. There will also be a section of CPAP monitoring where we shall consider the time of CPAP initiation, the mode of CPAP, the positive end expiratory pressure (PEEP) and complications during CPAP. The CRFs will also have a section on CPAP outcome, which will include the duration of CPAP, need for bag and mask ventilation, episodes of apnoea requiring stimulation, capillary refill more than 3 seconds, and episodes of bradycardia. SAS score will be recorded at CPAP initiation then 4-6 hours after initiation or as clinically indicated. Higher SAS scores will indicate worsening respiratory distress (Maximum of 10 scores).

CPAP monitoring and failure trigger forms will be filled to assess the neonates at risk of CPAP failure. There will be silverman – Anderson score sheet to assess the severity of respiratory distress and predict CPAP failure, CPAP monitoring and failure trigger form to track deterioration and identify early failure.

Tools used to collect data will be Case Report Forms (CRFs) specifically designed for this study to capture comprehensive neonatal, maternal, and clinical data. These forms will include information on gestational age, birth weight, gender, mode of delivery, APGAR scores, antenatal corticosteroids use, maternal infections and place of delivery. Clinical indicators such as Silverman-Anderson Score, FiO2 requirements and signs of respiratory distress will be recorded at baseline and every 4-6 hours after CPAP initiation. There will also be a section of CPAP monitoring where we shall consider the time of CPAP initiation, the mode of CPAP, the positive end expiratory pressure (PEEP) and complications during CPAP. The CRFs will also have a section on CPAP outcome, which will include the duration of CPAP, need for bag and mask ventilation, episodes of apnoea requiring stimulation, capillary refill more than 3 seconds, and episodes of bradycardia. SAS score will be recorded at CPAP initiation then 4-6 hours after initiation or as clinically indicated. Higher SAS scores will indicate worsening respiratory distress (Maximum of 10 scores). CPAP monitoring and failure trigger forms will be filled to assess the neonates at risk of CPAP failure. There will be Silverman – Anderson score sheet to assess the severity of respiratory distress and predict CPAP failure, CPAP monitoring and failure trigger form to track deterioration and identify early failure.

Neonates will be monitored for signs of CPAP failure using predefined criteria such as FiO2> 60% for more than one hour to maintain SPO2 90-95%, three or more apnea episodes requiring stimulation or any episodes requiring bag-mask ventilation in one hour, bradycardia (<100 bpm), capillary refill >3 seconds, persistent or worsening respiratory distress by SAS score. Outcomes including CPAP duration, referral for mechanical ventilation, length of NICU stay and final clinical outcome whether discharged, referral or death will be recorded. To ensure consistency and accuracy trained data collectors will follow standardized procedures and the Principal Investigator will conduct daily reviews for data quality and protocol adherence.

Neonates will be monitored for signs of CPAP failure using predefined criteria such as FiO2> 60% for more than one hour to maintain SPO2 90-95%, three or more apnea episodes requiring stimulation or any episodes requiring bag-mask ventilation in one hour, bradycardia (<100 bpm), capillary refill >3 seconds, persistent or worsening respiratory distress by SAS score. Outcomes including CPAP duration, referral for mechanical ventilation, length of NICU stay and final clinical outcome whether discharged, referral or death will be recorded. To ensure consistency and accuracy trained data collectors will follow standardized procedures and the Principal Investigator will conduct daily reviews for data quality and protocol adherence.

The final outcomes i.e. discharged from hospital; referral or death will be recorded during the monitoring period and at the end of the follow-up period. All data will be reviewed daily by the Principal Investigator and clinical supervisors to ensure completeness in data, accuracy and protocol adherence similar to the quality assurance measures used in other sub-Saharan neonatal studies.

## Data Availability

All data produced in the present study are available upon reasonable request to the authors

## Data management and analysis plan

The final outcomes i.e. *discharged from hospital; referral or death* will be recorded during monitoring period and at the end of the follow-up period of seven days of life.

All data will be reviewed daily by the Principal Investigator and supervisors to ensure completeness, accuracy and protocol adherence similar to the quality assurance measures used in other sub-Saharan neonatal studies. Besides, the PI will be performing data entry at each end of business day and captured data stored in the PI’s laptop until analysis time.

Exploratory data analysis will be performed prior to final analysis. Descriptive statistics will be summarized using means with standard deviations (if normally distributed) or medians with interquartile ranges (if skewed). The categorical variables, including gender and mode of delivery will be summarized as counts and percentages. These proportions will be stratified by gestational age and presented as cross-tabulations. To determine the magnitude of CPAP failure; a calculation of the proportion of neonates who meet predefined CPAP failure criteria during hospitalization will be made. This will be expressed as an equation; CPAP failure rates = number of neonates who failed CPAP/Total number of neonates on CPAP multiply by 100%. To provide a measure of precision we shall use the 95% confidence interval for this proportion. To explore the clinical predictors of failure, perform bivariate analysis will be used to compare neonates who failed to those who did not. The Chi-squared test or Fisher’s test will be used to assess the association between the categorical predictors (Mode of delivery, gender) and CPAP failure. Independent t-tests will be applied for the continuous variables (such as GA, Birth weight, FiO_2_ at initiation, and SAS score if the data is normally distributed. If the data will be skewed the the Mann-Whitney U tests will be applied. Variables demonstrating a p-value <0.20 in bivariate analysis will be included in a multivariable linear model to identify independent predictors of CPAP failure. Results will be reported as adjusted absolute risks with 95% confidence interval. Data analysis will be performed using SPSS version 26 (SPSS, IBM – NY, USA). All statistical analyses will be done assuming two-tailed statistics and an α-level of 5% will be used as a limit for type 1 error in findings.

## Ethics and Ethical considerations

Ethical clearance for the study will be sought from the Institutional Research and Ethics Committee of Kairuki university in Dar es Salaam, Tanzania. Permission to conduct the follow-ups at each hospital will be sought from the office of the hospital directors of each hospital. Written informed consent will be sought from each mother of newborns prior to inclusion into the study. The consenting process and the information on consent will include the goal of the study and its resultant follow-ups, risks and benefits associated with participating into the study, voluntary nature of participation to each newborn (and that refusal to participate will not inter alia interfere with the required services at the neonatal/hospital set ups, mothers of each recruited children allowed to ask questions and even given time to think about their response for participation as well as the allowance of mothers to retain contact address of both the PI and even the chair of the IRB (IREC-KU).

## AUTHORS’ CONTRIBUTIONS

RGS - Conception, Design & methods; funding and material resources, tool testing, software, writing the original manuscript, editing and approving the final protocol manuscript [PRINCIPAL INVESTIGATOR]

FSK – Conception, Design & Methods, supervision, editing and approving the final protocol manuscript. [CO-INVESTIGATOR]

MRF – Conception, Design & methods, supervision, editing and approving the final protocol Manuscript [CO-INVESTIGATOR]

NAD – Conception, Design & methods, editing and approving the final protocol manuscript [CO-INVESTIGATOR]

AIS - Conception, Design & methods, editing and approving the final protocol manuscript [CO-INVESTIGATOR]

KML – Conception, Design & methods, tools testing (reliability and validity), editing and approving the final protocol manuscript [CO-INVESTIGATOR]

## FUNDING STATEMENT

This research received no specific grant from any funding agency in the public, commercial or not-for-profit sectors

## COMPETING INTEREST STATEMENT

All authors declare NO COMPETING INTEREST exist in the preparation and publishing this protocol

## Notes

### Competing Interest Statement

The authors have declared no competing interest.

### Author Declarations

Name: Institutional Research Ethics Committee - Kairuki university

